# Validity of Markerless Three-dimensional Wrist and Finger Motion Analysis Using Deep Learning: A Comparison with Goniometric Measurement

**DOI:** 10.64898/2026.01.07.26343634

**Authors:** Ryo Sakai, Yukari Iwaya, Makoto Haraguchi, Masayuki Kanamori, Tomonari Sugano, Koshi Murata, Takashi Ryoke, Keiji Ishida, Yasutaka Kobayashi

## Abstract

**Background:** Markerless motion analysis using deep learning is attracting attention in the field of rehabilitation; however, the three-dimensional measurement accuracy in finger joints, which are prone to self-occlusion, has not been sufficiently validated. This study aimed to validate the accuracy of finger joint angle measurements obtained using a marker-less system based on DeepLabCut (DLC) and Anipose by comparing it with the clinical standard of goniometric measurements.

**Methods:** Forty-one healthy adults were recruited. Videos from ten participants were used for DLC training, whereas the remaining 31 served as the analysis subjects. Eight flexion movements (wrist, thumb, index, and middle finger) were recorded using five synchronized cameras. The DLC tracked 2D keypoints, and Anipose performed 3D reconstruction to calculate the angles. Goniometric measurements were performed simultaneously for comparison. The agreement between methods was evaluated using Bland–Altman analysis to identify fixed and proportional biases.

**Findings:** High agreement was observed between the angles estimated by marker-less analysis and goniometer measurements, and most data points were within the 95% limits of agreement. However, a significant proportional bias, where the error increased with an increase in flexion angle, was observed in distal joints, such as the thumb interphalangeal joint and index/middle proximal interphalangeal joints.

**Interpretation:** This system demonstrated clinically acceptable validity for measuring finger range of motion. However, underestimation is likely to occur in the distal finger joints and at maximal flexion owing to the influence of occlusion, necessitating consideration of proportional bias. This method represents a noninvasive, low-cost tool for assessing hand function.

## 1. Introduction

Markerless motion capture (MMC) technology has advanced rapidly in recent years, and pose estimation methods that apply deep learning have attracted attention in the field of rehabilitation medicine (Needham et al., 2021). Comprehensive overviews have been published regarding the principles and potential of deep-learning-based motion capture, including advantages such as flexible customizability and noninvasiveness (Mathis et al., 2020; Mathis and Mathis, 2020). DeepLabCut (DLC) is a framework that precisely tracks specific body parts based on labeled training data provided on images and is widely utilized in animal research and human movement analysis (Mathis et al., 2018). Moreover, the utility of deep learning tools such as DLC has been emphasized in neuroscience and behavioral measurements (Mathis and Mathis, 2020). Anipose (Karashchuk et al., 2021), which reconstructs three-dimensional posture from multiview videos, integrates two-dimensional keypoints estimated by DLC, enabling markerless 3D motion analysis. These deep learning-based markerless methods have multiple advantages over conventional marker-based three-dimensional motion analysis, such as (1) reduced burden on the body, (2) expected reduction of inter-rater error, and (3) ease of operation in clinical settings (Chatzis et al., 2020; Maggioni et al., 2025). However, studies utilizing the MMC still lack evaluation in patient populations, and the necessity of validity verification has been highlighted (Pardell et al., 2024). Deep learning-based MMC has been applied to upper limb movement evaluation in stroke survivors, with confirmed correlations with clinical scores (Lam et al., 2025), highlighting the potential of this technology for fine motor evaluation of the fingers.

In particular, fingers contain closely packed small joints, making them prone to self-occlusion during flexion or grasping movements (Chatzis et al., 2020), and because of the structural characteristics of short link lengths where joint angles depend on minute coordinate variations, they are reported to be susceptible to depth bias during 3D reconstruction (Lecomte et al., 2021; Zimmermann et al., 2017; Ge et al., 2019). This depth error issue is particularly pronounced in research estimating 3D poses from a single RGB image in highly articulated body regions with complex structures, such as fingers (Zimmermann et al., 2017). Furthermore, many previous studies using DLC targeted the entire upper limb or gross postural changes, with very few analyses focusing on fine motor movements of the fingers (Gionfrida et al., 2022). Considering the geometric constraints unique to fingers (short link lengths, small range of motion, and poor visibility of distal joints), verifying the validity of marker-less methods for 3D finger motion analysis is a crucial task for clinical application and research utilization. However, to the best of our knowledge, no study has been found that directly compared the estimation of 3D wrist and finger joint angles using DLC and Anipose combined with a clinical standard measurement such as a goniometer. The objective of this study was to clarify the validity of wrist and finger joint angle estimation using these markerless deep learning systems through a comparison with goniometric measurements (Fig.1).

**Figure 1.**
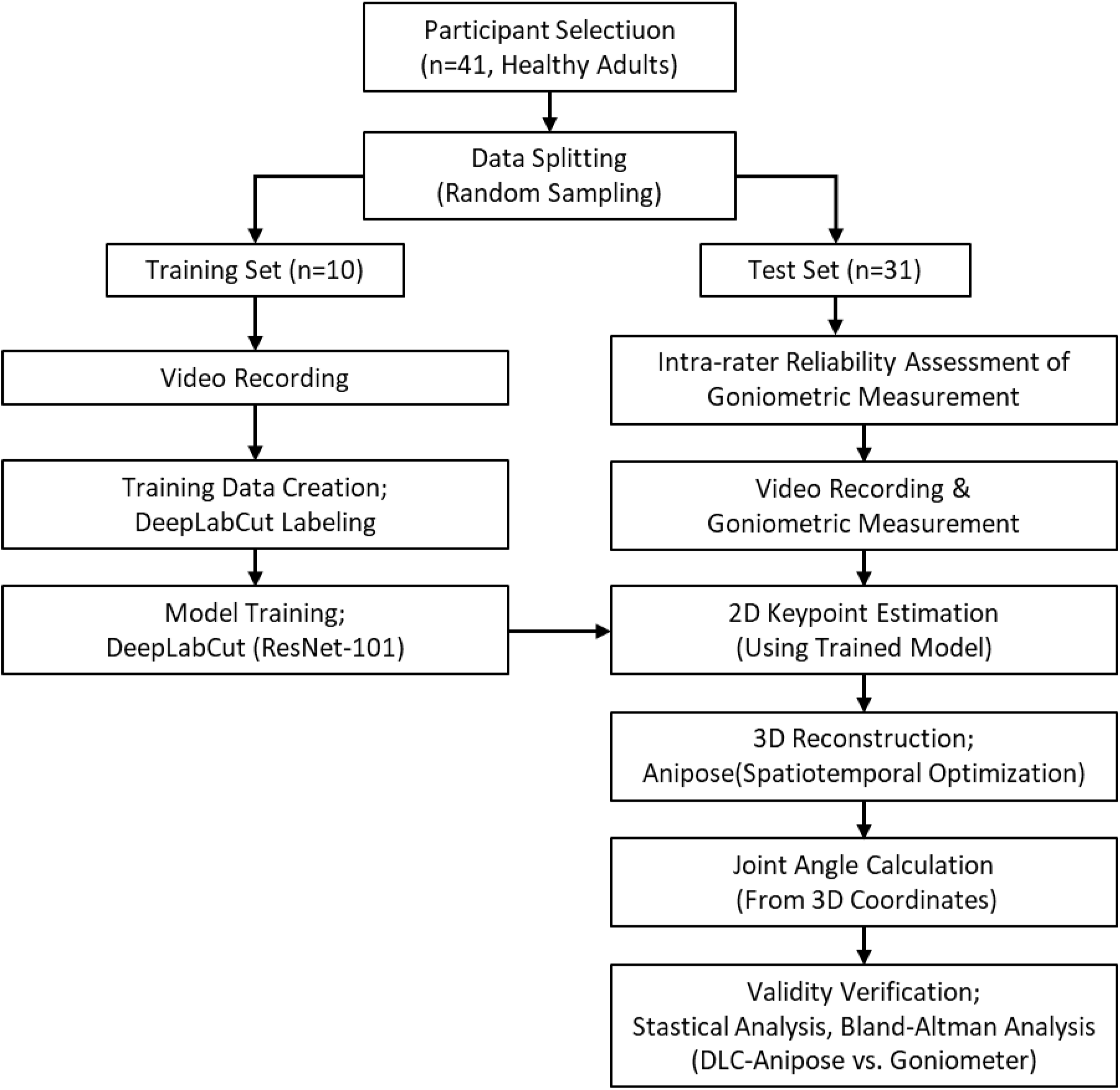
Overview of the Data Acquisition and Analysis Procedure for Markerless 3D Wrist and Finger Motion Analysis.

## 2. Methods

### 2.1 Participants

Forty-one healthy adults participated in this study. Individuals with a history of orthopedic or neurological conditions affecting the upper limb or significant finger deformities were excluded. Videos from 10 randomly extracted participants were used to create the DLC training data, and the remaining 31 participants’ videos, excluding these 10, were used for analysis. The training data group consisted of 7 females and 3 males (mean age 21.1±0.3 years), and the analysis data group consisted of 24 females and 7 males (mean age 20.4±0.5 years). All participants were informed of the research purpose and procedures, and written consent was obtained from all participants. This study was approved by the Ethical Review Committee of Nittazuka Medical Welfare Center (approval no. Nittazuka Ethics 2024–12).

### 2.2 Experimental Setup

All measurements targeted the right upper limb and the participants maintained a seated posture with their forearms naturally placed on a table embedded in a transparent acrylic plate (Fig. 2A). This posture was adopted because it allowed stable observation of the fingers from multiple viewpoints and did not impede maximum flexion movements. The participants were instructed to slowly perform eight movements: wrist flexion/extension, thumb metacarpophalangeal (MCP)/interphalangeal (IP) joint flexion, index finger MCP/proximal interphalangeal (PIP) joint flexion, and middle finger MCP/PIP joint flexion from a neutral intermediate position to the maximum flexion position, and to hold the maximal flexion position (Fig. 2B). The left upper limb was placed alongside the body or thigh to avoid camera capturing.

**Figure 2.**
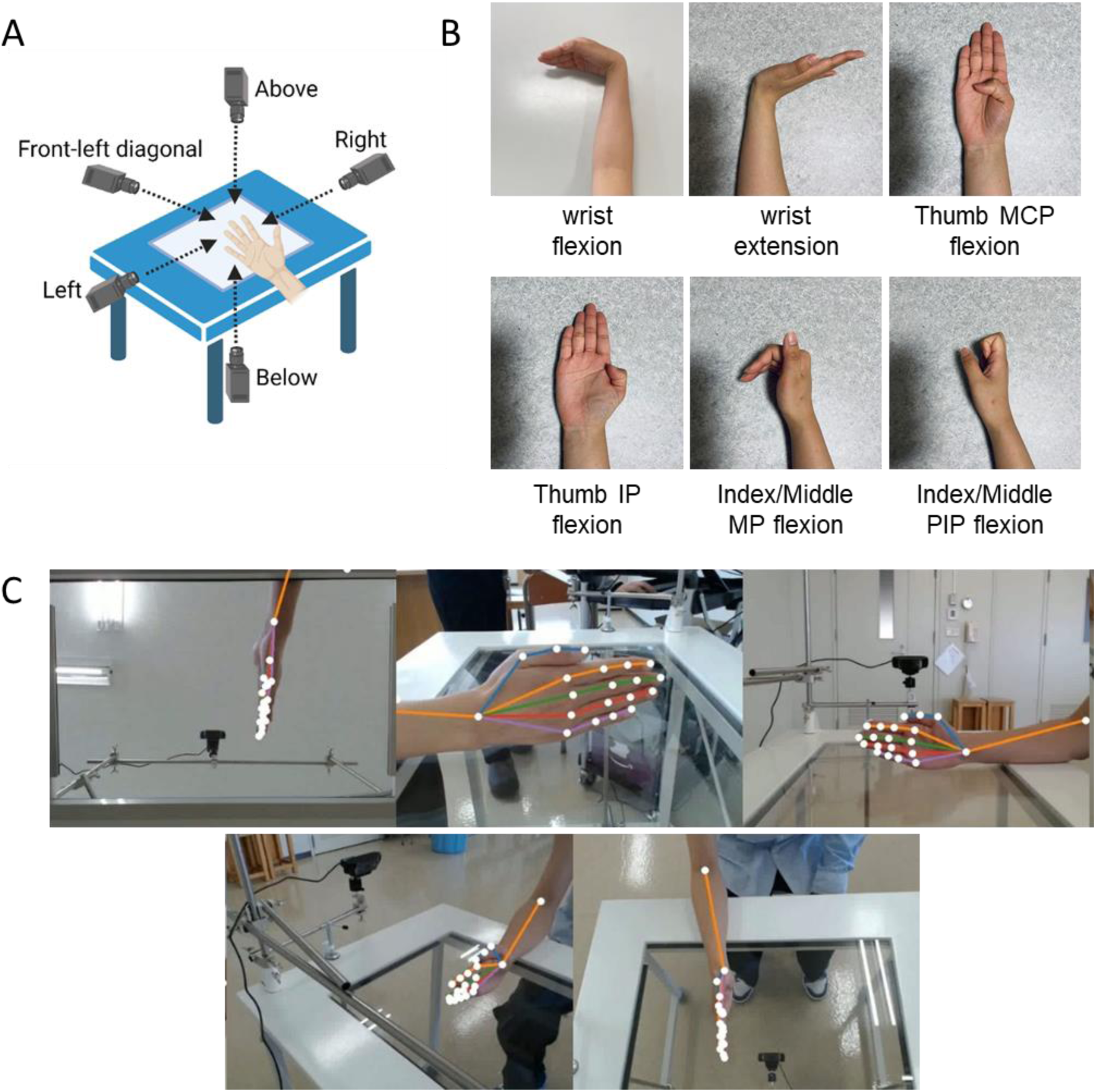
Example of Measurement Setup and Markerless Analysis (DLC/Anipose) A: Filming environment. Participant posture and camera placement. The participant was seated with the forearm placed naturally on a clear acrylic plate, and five cameras were positioned for recording below, laterally left/right, frontally, left anteriorly, and above. B: Measurement posture. C: Example of keypoint tracking using DLC-Anipose.

### 2.3 Video recording

Five video cameras (C922n, Logicool Co. Ltd., Tokyo, Japan) were used to record the movements positioned below, to the left and right, in front, front-left, and above. This created an environment in which parts prone to occlusion, such as the interphalangeal joints, could be observed in multiple directions. All cameras filmed in Full HD (1920×1080 pixels) at 60 fps. The start and end of the recording were synchronized using OBS Studio, and the video from each camera was output in MP4 format.

### 2.4 DeepLabCut Processing

DLC (version 2.2) was used for the automated extraction of the finger joint points (Mathis et al., 2018; Nath et al., 2019). In this study, to select frames including diversity in finger shapes and occlusion, an automatic extraction function using DLC’s k-means clustering of DLC was used, and 250 frames (five frames per camera from 10 participants, across five cameras) were adopted as training data. The extracted frames were unified into 640×480 pixels. Twenty-one keypoints required to describe hand motion (elbow, wrist, thumb MCP joint, IP joint, and fingertip), and the MCP, PIP, distal interphalangeal (DIP) joints, and fingertip of the index to little fingers were defined and manually labeled by a rater who did not perform goniometric measurements. A standard DLC network with ResNet-101 as the backbone was used for the training model, and weights pretrained on ImageNet were used as the initial values. The dataset was divided into 95% for training and 5% for testing. For the training settings, stochastic gradient descent was used as the optimization algorithm, and 500,000 learning iterations were executed according to the DLC’s multi-step learning rate policy. To prevent overfitting, multiple data augmentation techniques enabled by default in DLC (such as rotation±25°, random crop, scale jitter, CLAHE, histogram equalization, sharpening, and embossing) were applied. A batch size of one was set for the training phase and eight for the inference phase. The accuracy of the learned model was evaluated using the root-mean-square error (RMSE) against the test data, and the RMSE of all keypoints was calculated. During the analysis, keypoints with a likelihood estimated by DLC of less than 0.6 were excluded or interpolated to suppress error propagation due to unreliable coordinates.

### 2.5 Computing Environment

All the training and analysis processes were executed using a workstation (Dell Precision 7920 Tower; CPU: Dual Intel Xeon Gold 6134 @ 3.20GHz, RAM: 256 GB; GPU: NVIDIA GeForce GTX 1080 8GB) (Mathis et al., 2018).

### 2.6 Anipose Calibration and 3D Reconstruction

The Anipose library was used for 3D motion analysis (Karashchuk et al., 2021). Camera calibration was performed using a Charuco board (10×7 grid; square size: 25 mm; marker size: 18.75 mm). Two hundred to 250 frames of detection data were acquired for each camera, and the intrinsic and extrinsic parameters were estimated using the fisheye model to correct lens distortion. The mean re-projection error was 0.39 pixels, confirming sufficient accuracy for 3D reconstruction. For 3D reconstruction, Anipose’s spatiotemporal optimization method was applied. This method minimizes the reprojection error by imposing the consistency of the limb length as a constraint instead of RANSAC by integrating information from all camera viewpoints. The threshold for the re-projection error was set to three pixels. For the reconstructed 3D trajectories, spatiotemporal smoothing (spatial smoothness: 25; temporal consistency weight: 10), performed internally by Anipose, was applied to reduce coordinate noise.

### 2.7 Joint Angle Computation

Joint angles were calculated based on the reconstructed 3D coordinates (Fig. 2C). The flexion angle of each joint was defined as the angle in the 3D space formed by the proximal and distal segment vectors (e.g., the MCP-PIP and PIP-DIP vectors) (Needham et al., 2021). The maximum flexion angle for each movement was extracted from the calculated time-series data and used for statistical analysis.

### 2.8 Reference Measurement

Goniometric measurements obtained as reference values were consistently performed by an occupational therapy student under the guidance and supervision of an experienced occupational therapist. To verify the reliability of the range of motion measurement for the wrist and finger joints, wrist flexion and index PIP flexion were measured three times for all 31 subjects as representatives of each joint, and intra-rater reliability was calculated using the intraclass correlation coefficient (ICC[1,3]) (Koo & Li, 2016). For each trial, the subject was instructed to return the limb to a relaxed angle once and then assume the designated posture again. The goniometers were blinded to the DLC analysis results. These procedures prevented measurement bias and ensured that each measurement result was obtained independently. To measure the actual tasks, the subject was instructed to pause at each posture during video recording, and the joint angles were measured after securing the timing for analysis.

### 2.9 Statistical Analysis

This study was planned as an exploratory method comparison study targeting a sample size of 30 or more, referring to the sample sizes in previous studies focusing on range of motion measurement and motion capture (generally 20–40 participants) (Needham et al., 2021; Lam et al., 2025; Nakano et al., 2020). The agreement between DLC/anipose three-dimensional angles and goniometric measurements was evaluated by Bland–Altman analysis (Bland & Altman, 1986; Bland & Altman, 1999). A one-sample t-test was used to examine the presence of fixed bias, and a simple regression analysis with the difference as the dependent variable and the mean as the independent variable was used to evaluate proportional bias (proportional bias). The 95% limits of agreement (LOA) were calculated to verify the clinical agreement. SPSS (version 23.0, IBM Corp., Armonk, NY, USA) and GraphPad Prism 8 (GraphPad Software Inc., Boston, MA, USA) were used for the statistical analyses. All statistical tests were two-sided, and the significance level was set at *P* < 0.05.

## 3. Results

### 3.1 Reliability of the reference measurement

Table 2 shows the intrarater reliability of the goniometric measurements used as reference measurements. In the measurements obtained from the participants, the ICC[1,3] for wrist flexion was 0.985, and that for index PIP joint flexion was 0.958, indicating excellent reliability (ICC > 0.95) for both. The standard errors of measurement (SEM) was 1.42° for the wrist joint and 1.04° for the index finger PIP joint, and the minimal detectable changes (MDC) was 3.93° and 2.89°, respectively. This confirms that the goniometric measurements in this study were sufficiently reproducible for comparison.

**Table 1.** Participant characteristics.

|  | Training set | Test set |
| --- | --- | --- |
| Number of participants | 10 | 31 |
| Age (years), mean ± SD | 21.1 ± 0.3 | 20.4 ± 0.5 |
| Sex (women/men) | 7 / 3 | 24 / 7 |
| Used for | DLC training | 3D analysis |

SD; standard deviation, DLC; DeepLabCut.

**Table 2.** Intra-rater reliability of goniometric measurements.

| Joint / Movement | Mean±SD<br>(°) | ICC[1,3] | *P* value | 95% CI | SEM | MDC |
| --- | --- | --- | --- | --- | --- | --- |
| Wrist flexion | 76.2±11.6 | 0.985 | <0.001 | 0.973-0.992 | 1.416 | 3.926 |
| Index PIP flexion | 97.9±5.1 | 0.958 | <0.001 | 0.924-0.978 | 1.042 | 2.889 |

SD, standard deviation; ICC, intraclass correlation coefficient; CI, confidence interval; SEM, standard error of measurement; MDC, minimal detectable change; PIP, proximal interphalangeal joint.

### 3.2 Agreement between Markerless 3D Estimates and Goniometer Measurements

#### 3.2.1 Fixed bias

The agreement between the estimates by the DLC/anipose system and goniometer measurements was examined using Bland–Altman analysis (Fig. 3A-H, Table 3). At the wrist joint, positive mean differences of 10.86 ± 10.73° for flexion and 6.79 ± 6.14° for extension were observed, both indicating a fixed bias where the system overestimated the angle (both *P* < 0.0001). Conversely, thumb MCP flexion showed a large negative mean difference of −15.85 ± 10.69°, indicating a consistent underestimation by the estimated value (*P* < 0.0001). At the fingers’ MCP and PIP joints, the mean differences were relatively small, and no fixed bias was observed for index MCP flexion (3.67 ± 11.42°, *P* = 0.084) and index PIP flexion (0.29 ± 12.35°, *P* = 0.897). Middle MCP flexion (3.86 ± 14.28°, *P* = 0.143) also did not show significant fixed bias, but middle PIP flexion showed a gentle negative mean difference of −8.83 ± 21.76°, which was detected as significant fixed bias (*P* = 0.031). Thumb IP flexion showed almost no difference, −0.06 ± 16.14° (*P* = 0.983), and fixed bias was absent.

**Figure 3.**
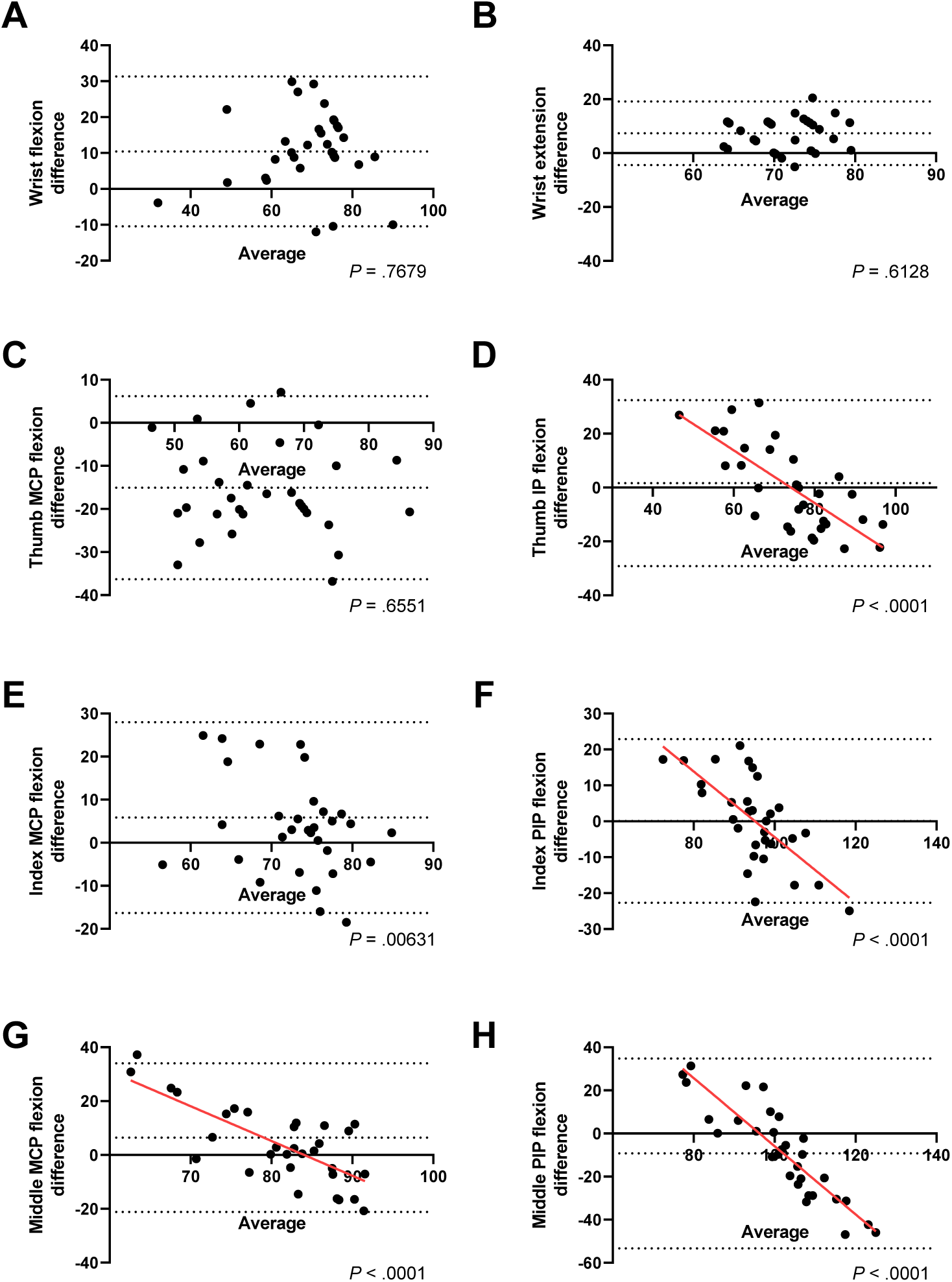
Agreement of Eight Joint Motions Assessed Using Bland–Altman Plots Bland–Altman plots showing the difference between the estimated angles by DLC/Anipose and the goniometric measurements (vertical axis) versus the mean value of both methods (horizontal axis). Each panel corresponds to wrist flexion/extension, and MCP/IP/PIP flexion of the thumb, index, and middle fingers. The central dashed line represents the mean difference (fixed bias), and the upper and lower dashed lines indicate the 95% limits of agreement (LOA). Joints for which significant proportional bias was detected show a significant negative slope of the regression line (red solid line).

**Table 3.**
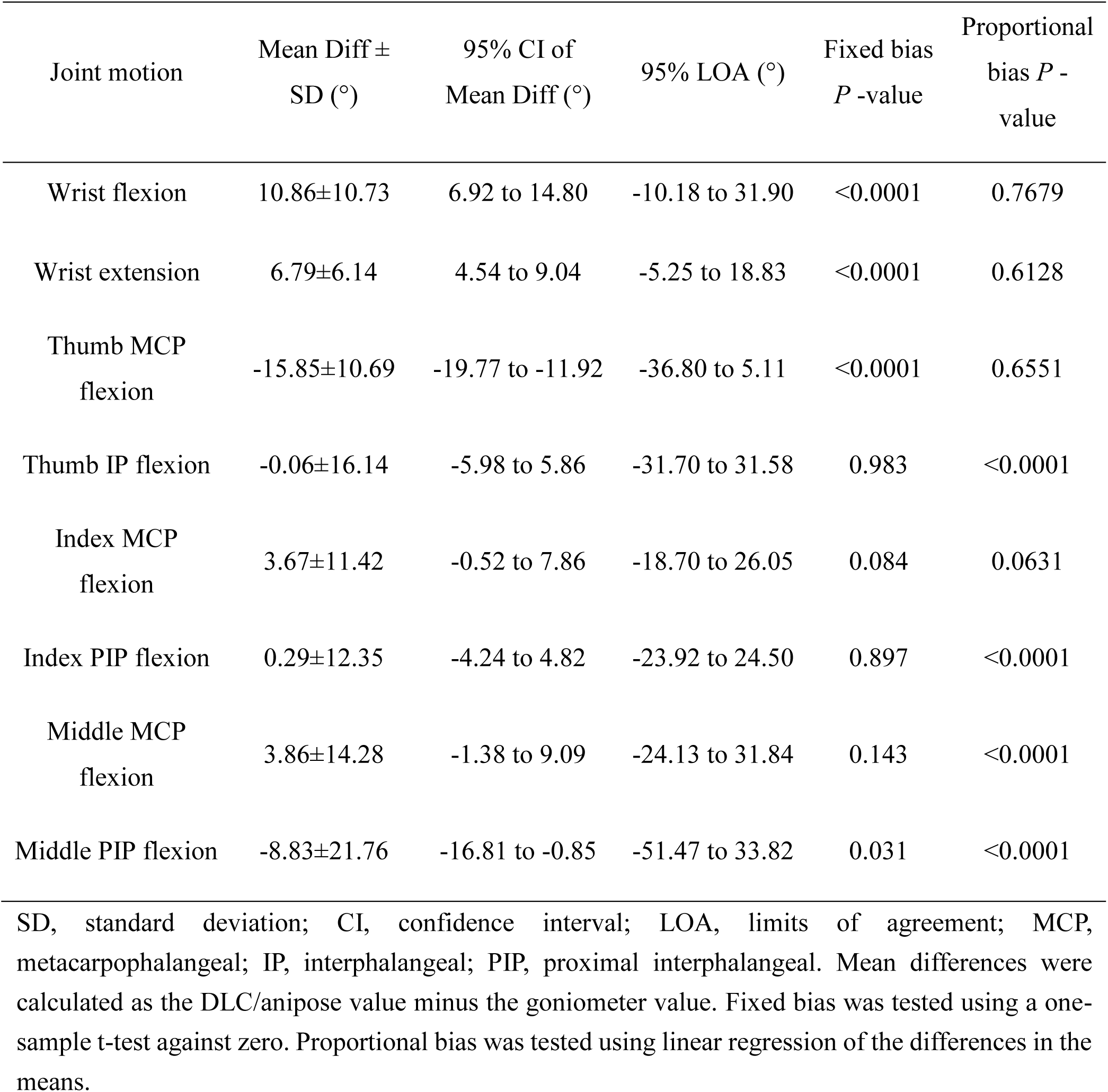
Bland–Altman analysis results.

| Joint motion | Mean Diff ±<br>SD (°) | 95% CI of<br>Mean Diff (°) | 95% LOA (°) | Fixed bias<br>*P*-value | Proportional<br>bias *P*-<br>value |
| --- | --- | --- | --- | --- | --- |
| Wrist flexion | 10.86±10.73 | 6.92 to 14.80 | -10.18 to 31.90 | <0.0001 | 0.7679 |
| Wrist extension | 6.79±6.14 | 4.54 to 9.04 | -5.25 to 18.83 | <0.0001 | 0.6128 |
| Thumb MCP<br>flexion | -15.85±10.69 | -19.77 to -11.92 | -36.80 to 5.11 | <0.0001 | 0.6551 |
| Thumb IP flexion | -0.06±16.14 | -5.98 to 5.86 | -31.70 to 31.58 | 0.983 | <0.0001 |
| Index MCP<br>flexion | 3.67±11.42 | -0.52 to 7.86 | -18.70 to 26.05 | 0.084 | 0.0631 |
| Index PIP flexion | 0.29±12.35 | -4.24 to 4.82 | -23.92 to 24.50 | 0.897 | <0.0001 |
| Middle MCP<br>flexion | 3.86±14.28 | -1.38 to 9.09 | -24.13 to 31.84 | 0.143 | <0.0001 |
| Middle PIP flexion | -8.83±21.76 | -16.81 to -0.85 | -51.47 to 33.82 | 0.031 | <0.0001 |

SD, standard deviation; CI, confidence interval; LOA, limits of agreement; MCP, metacarpophalangeal; IP, interphalangeal; PIP, proximal interphalangeal. Mean differences were calculated as the DLC/anipose value minus the goniometer value. Fixed bias was tested using a one-sample t-test against zero. Proportional bias was tested using linear regression of the differences in the means.

#### 3.2.2 Limits of Agreement

The 95% LOA varied significantly depending on the joint, and the spread of the error distribution was also dependent on the joint characteristics. While wrist flexion had a relatively narrow range of −10.18 to 31.90°, middle PIP flexion showed the widest range of −51.47 to 33.82°, suggesting a tendency for larger error variability in distal joints.

#### 3.2.3 Proportional bias

Regression analysis was performed to verify whether the estimation error depended on the magnitude of the angle; the slope was not significant for wrist flexion (*P* = 0.7679), wrist extension (*P* = 0.6128), thumb MCP flexion (*P* = 0.6551), and index MCP flexion (*P* = 0.0631), and proportional bias was not observed. In contrast, a significant proportional bias was detected for the thumb IP flexion (*P* < 0.0001), index PIP flexion (*P* < 0.0001), middle MCP flexion (*P* < 0.0001), and middle PIP flexion (*P* < 0.0001), all of which showed negative slopes. In particular, middle PIP flexion showed the largest slope, and the tendency for underestimation (where the estimated value became smaller than the measured value) to increase in an angle-dependent manner as the flexion angle increased was pronounced.

## 4. Discussion

The purpose of this study was to verify the validity of markerless three-dimensional finger motion analysis using DLC and Anipose compared to the clinical standard goniometer. The fixed and proportional biases were small in the proximal joints, such as the wrist joint, suggesting that this system possesses clinically acceptable accuracy. This is consistent with previous studies suggesting that conventional marker-less analysis is highly adaptable to gross joint movements (Nakano et al., 2020). Research using DLC reports a very high agreement in sagittal plane kinematics during vertical jumps (RMSE<3.22°) (Drazan et al., 2021), and the results of this study also support the idea that marker-less tracking can identify the target site with high accuracy. Furthermore, as the reference goniometer measurement showed high intra-rater reliability (ICC > 0.95), the reliability of this comparative validation was confirmed. However, a pronounced proportional bias was confirmed in the distal joints (IP and PIP), where the measurement error increased with an increase in flexion angle. The negative slope in the regression analysis of the Bland–Altman plot indicates a tendency of the system to underestimate the actual angle as the flexion angle increases. The primary cause is thought to be the influence of self-occlusion and foreshortening that occurs during maximal flexion of the fingers (Chatzis et al., 2020; Zimmermann et al., 2017). The accuracy of deep learning models tends to decrease when the training dataset is labeled inaccurately without considering biomechanical applications or when extreme changes in shape are not included in the training data (Needham et al., 2021; Wade et al., 2022). Furthermore, the 3D reconstruction process by anipose was also an error factor. Lecomte et al. (2021) reported that when the object is small relative to the camera baseline, the estimation error in the depth direction is amplified (Lecomte et al., 2021). In fact, previous studies have shown that conventional marker-less devices, such as the Leap Motion Controller, exhibit large errors, particularly in spatial and temporal accuracy, compared to criterion systems (Niechwiej-Szwedo et al., 2018), suggesting that securing accuracy in fine motor analysis of the fingers has been a long-standing challenge. Although multiview optimization was used in this study, in addition to the challenges of joints with short link lengths, such as the fingers, the estimation of 3D rotation angles of large lower extremity joints is also reported to have large errors compared to marker-based systems (D’Souza et al., 2024; Kanko et al., 2021), which supports the claim that the problem of rotational and depth errors in 3D space in this study is particularly exacerbated in the fingers owing to their short segments. Furthermore, the reliability evaluation of AI-driven MMC reported high reliability in the sagittal plane but low reliability in the frontal and transverse planes, which is consistent with the findings of this study (Schoenwether et al., 2025). The maximal error observed, particularly at the middle PIP joint, was likely due to its central position in the palm, making it susceptible to occlusion from all camera viewpoints, thus increasing the uncertainty of 3D reconstruction. Despite these limitations, the high agreement observed at the wrist and MCP joint levels suggests that this system is useful for screening and gross range of motion assessment (Needham et al., 2021; Gionfrida et al., 2022; Lam et al., 2025). Previous research using conventional MMC technology has validated its applicability to hand kinematic measurements in upper limb rehabilitation (Metcalf et al., 2013), indicating that hand motion analysis methods have long been explored for applying markerless approaches to clinical settings. This method, capable of measurement using a non-contact and low-cost setup, possesses clinical advantages not found in conventional contact goniometers, such as the ability to avoid pain or infection risks associated with contact and high versatility in freely setting measurement points (Maggioni et al., 2025). This study had several limitations. First, because this was an exploratory investigation, convenience sampling was used, and there was a possibility of bias in the sample size and participant attributes. Secondly, the sample size was limited to healthy adults. In patients with finger deformities owing to rheumatoid arthritis or contracture, the skin texture and skeletal structure differ from those of the standard model, potentially leading to a lower accuracy than that obtained in this study. Third, static-holding posture measurements were adopted to ensure a stable comparison. Further accuracy validation is required in dynamic situations, such as actual clinical movements, as motion blurring and more complex occlusions occur. In conclusion, although this system is a promising tool for finger motion analysis, it is necessary to note that proportional bias occurs in the measurements of the distal joints and at maximum flexion. Future directions, such as augmenting diverse training data corresponding to deformed hands and implementing transformer-based models (Zheng et al., 2021) that integrate time-series information in a more sophisticated manner, will be key to improving the accuracy of clinical applications.

## 5. Conclusions

The validity of the markerless 3D finger motion analysis using DLC and Anipose was confirmed by comparison with goniometric measurements. This method can be a useful tool for clinical research and rehabilitation assessments under appropriate visual conditions and analytical settings. It is expected that this will contribute to new assistance in hand function evaluation through further improvement in analysis accuracy and the development of finger-specific models in the future.

## CRediT authorship contribution statement

Ryo Sakai: Conceptualization, Data curation, formal analysis, funding acquisition, Investigation, Methodology, Project administration, Software, writing the original draft, and visualization. Yukari Iwaya: Methodology, Investigation, Validation, Writing, review, and editing. Makoto Haraguchi: Methodology, Resources, Writing, review, and editing. Masayuki Kanamori: Methodology, Writing – Review, and Editing. Tomonari Sugano: Methodology, Writing, review, and editing. Koshi Murata: Methodology, Writing – Review, and Editing. Takashi Ryoke: Methodology, Writing, review and editing. Keiji Ishida: Conceptualization, Methodology. Yasutaka Kobayashi: Project administration, Resources, Supervision, Writing, review, and editing.

## Acknowledgments

Figures were created using Bio-Render.

## Data availability

Data will be made available on reasonable request.

